# Sex differences in risk factors, burden, and outcomes of cerebrovascular disease in Alzheimer’s disease populations

**DOI:** 10.1101/2023.02.13.23285819

**Authors:** Cassandra Morrison, Mahsa Dadar, D. Louis Collins, the Alzheimer’s Disease Neuroimaging Initiative

**Affiliations:** McConnell Brain Imaging Centre, Montreal Neurological Institute, McGill University, Montreal, Quebec, Canada; Department of Neurology and Neurosurgery, McGill University, Montreal, Quebec, Canada; Department of Psychiatry, McGill University, Montreal, Quebec, Canada; Douglas Mental Health University Institute, Montreal, Quebec, Canada

**Author notes:** **Corresponding author:** Cassandra Morrison, Montreal Neurological Institute, 3801 University Street, Montreal QC, H3A 2B4. Data used in preparation of this article were obtained from the Alzheimer’s Disease Neuroimaging Initiative (ADNI) database (adni.loni.usc.edu). As such, the investigators within the ADNI contributed to the design and implementation of ADNI and/or provided data but did not participate in analysis or writing of this report. A complete listing of ADNI investigators can be found at: http://adni.loni.usc.edu/wp-content/uploads/how_to_apply/ADNI_Acknowledgement_List.pdf.

**Keywords:** Older Adults, Sex Differences, White Matter Hyperintensities

## Abstract

**Background:** White matter hyperintensity (WMH) accumulation is associated with vascular risk factors such as hypertension, diabetes, smoking, and obesity. Increased WMH burden results in increased cognitive decline and progression to mild cognitive impairment (MCI) and dementia. However, research has been inconsistent when examining whether sex differences influence the relationship between vascular risk factors, WMH accumulation, and cognition.

**Methods:** A total of 2119 participants (987 females) with 9847 follow-ups from the Alzheimer’s Disease Neuroimaging Initiative met inclusion criteria for this study. Linear regressions were used to examine the association between vascular risk factors (individually and as a composite score) and WMH burden in males and females. When controlling for vascular risk factors, linear mixed models were also used to investigate whether the relationship between WMHs and longitudinal cognitive scores differed between males and females.

**Results:** Males had overall increased occipital (*p*<.001), but lower frontal (*p*<.001), total (*p*=.01), and deep (*p*<.001) WMH burden compared to females. For males, history of hypertension was the strongest contributor to WMH burden. On the other hand, the vascular composite score was the strongest factor for WMH in females. Greater increase in WMH accumulation was observed in males with a history of hypertension in the frontal region (*p*=.014) and males with high systolic blood pressure in the occipital region (*p*=.029) compared to females. With respect to cognition, WMH burden was more strongly associated with longitudinal decreases in global cognition, executive functioning, and functional activities of daily living in females compared to males.

**Discussion:** These findings show that controlling hypertension is important to reduce WMH burden in males. Conversely, minimizing WMH burden through vascular risk factors requires controlling many factors for females (e.g., hypertension, diabetes, smoking, alcohol abuse, etc.). The results have implications for therapies and interventions designed to target cerebrovascular pathology and the subsequent cognitive decline.

## Introduction

Evidence for cerebral small vessel disease (CSVD) can appear as white matter hyperintensities (WMHs) observed on T2-weighted or FLAIR magnetic resonance images (MRI). Some CSVD lesions can also be detected as white matter lesions (WMLs) on T1-weighted MRIs. Although WMHs are commonly observed in cognitively unimpaired older adults^1^ their presence has been linked to cognitive deterioration in healthy older adults^2^, and increased risk for progression to mild cognitive impairment (MCI)^3^ and dementia^4^. The presence and accumulation of CSVD and is associated with various vascular risk factors such as hypertension, diabetes, high body mass index (BMI), tobacco smoking, and alcohol consumption^5–10^. These cerebrovascular risk factors are typically thought to affect men more than women; however, after menopause women may be at a higher risk than men^11^. Furthermore, these risk factors are known to affect women differently than men^12^. For example, although men tend to have increased blood pressure early in life, women have a steeper increase in blood pressure that remains higher and often less controlled than men in mid life^12,13^. Risk factors exclusive to women such as gestational diabetes and preeclampsia are also associated with accelerated development of cerebrovascular disease^14^.

Sex related differences in these risk factors may influence why some studies have reported sex differences in the prevalence of WMHs. Whether differences exist between males and females in WMH burden has been inconsistent. Some studies have reported greater WMH burden in females than males^15–18^. Some have reported no sex differences in WMH burden^10,19^, while others have reported increased WMH burden in males^20^. In addition, several studies have noted that vascular risk factors such as hypertension are more strongly associated with WMH development in males than females^15,21,22^, whereas BMI has shown an association with increased WMH for both males and females^23^ and smoking with higher WMH in females^15^.

Examining sex differences in WMH has implications for the treatment and mitigation of cognition decline. Many of the underlying vascular risk factors associated with WMHs are potentially treatable or modifiable (e.g., hypertension, body mass index, smoking, alcohol consumption, type II diabetes). Therefore, identifying which risk factors are most likely to result in the accumulation of WMHs could help reduce their impact on cognition and the subsequent conversion to dementia may be mitigated. While previous research has observed that sex differences are present in WMH burden and in the association between WMHs and vascular risk factors the findings are conflicting and limited in number.

Given the lack of research on sex differences and WMHs it is possible that different risk factors are associated with WMH accumulation in males and females. The goal of this paper was thus to determine if sex:1) influences progression in WMH, 2) influences which vascular risk factors are associated with WMH burden, 3) interacts with risk factors to influence WMH burden, and 4) influences the relationship between WMH burden and cognition. This study will advance the current understanding of how sex influences pathological changes in aging and cognitive decline. Women tend to remain underrepresented in most research studies, with researchers failing to analyze by sex, biasing results and limiting generalizability^24^. Inclusion of studies examining sex differences is essential for this research to improve our knowledge about sex-specific influences on aging and dementia and to design targeted intervention strategies that apply to more representative samples.

## Methods

### Alzheimer’s Disease Neuroimaging Initiative

Data used in the preparation of this article were obtained from the Alzheimer’s Disease Neuroimaging Initiative (ADNI) database (adni.loni.usc.edu). The ADNI was launched in 2003 as a public-private partnership, led by Principal Investigator Michael W. Weiner, MD. The primary goal of ADNI has been to test whether serial MRI, positron emission tomography (PET), other biological markers, and clinical and neuropsychological assessment can be combined to measure the progression of mild cognitive impairment and early AD. The study received ethical approval from the review boards of all participating institutions. Written informed consent was obtained from participants or their study partner. Participants were selected only from all ADNI Cohorts (ADNI-1, ADNI-GO, ADNI-2 and ADNI-3).

### Participants

Full participant inclusion and exclusion criteria are available at www.adni-info.org. All participants were between the ages of 55 and 90 at baseline, with no evidence of depression. Cognitively healthy older adults exhibited no evidence of memory decline, as measured by the Wechsler Memory Scale and no evidence of impaired global cognition as measured by the Mini Mental Status Examination (MMSE) or Clinical Dementia Rating (CDR). MCI participants scored between 24 and 30 on the MMSE, 0.5 on the CDR, and abnormal scores on the Wechsler Memory Scale. Dementia was defined as participants who had abnormal memory function on the Wechsler Memory Scale, an MMSE score between 20 and 26 and a CDR of 0.5 or 1.0 and a probable AD clinical diagnosis according to the National Institute of Neurological and Communicative Disorders and Stroke and the Alzheimer’s Disease and Related Disorders Association criteria^25^.

Participants were included if they had MRIs from which WMHs could be extracted, Hachinski score, as well as completed information for all vascular risk factors (i.e., BMI, blood pressure, hypertension, diabetes, stroke, smoking, alcohol abuse).

### Structural MRI acquisition and processing

All scans were downloaded from the ADNI website (see http://adni.loni.usc.edu/methods/mri-tool/mri-analysis/ for the detailed MRI acquisition protocol). T1w scans for each participant were pre-processed through our standard pipeline including noise reduction^26^, intensity inhomogeneity correction^27^, and intensity normalization into range [0-100]. The pre-processed images were then linearly (9 parameters: 3 translation, 3 rotation, and 3 scaling) ^28^ registered to the MNI-ICBM152-2009c average^29^.

### WMH Measurements

A previously validated WMH segmentation technique was employed to generate participant WMH measurements^4^. This technique has been validated in ADNI in which a library of manual segmentations based on 50 ADNI participants (independent of those studied here) was created. The technique has also been validated in other multi-center studies such as the Parkinson’s Markers Initiative^30^ and the National Alzheimer’s Coordinating Center^31^. WMHs were automatically segmented using the T1w contrasts, along with a set of location and intensity features obtained from a library of manually segmented scans in combination with a random forest classifier to detect the WMHs in new images^32,33^. WMH load was defined as the volume of all voxels as WMH in the standard space (in mm^3^) and are normalized for head size. The volumes of the WMHs for frontal, parietal, temporal, and occipital lobes as well as the entire brain were calculated based on regional masks from the Hammers atlas^32,34^. The quality of the registrations and WMH segmentations was visually verified by an experienced rater (author M.D.), blinded to diagnostic group.

In addition, Deep and Periventricular (PV) WMHs were also obtained through a previously validated technique that started by segmenting the ventricles on the T1w images with a patch-based label fusion segmentation technique^35,36^. People with AD were included in the training library to ensure that this technique could accurately segment the larger ventricles observed in people with AD. All ventricle segmentations were visually inspected (by Author M.D.), those that did not pass quality control were excluded from the PV and deep WMH analyses (N=9). The ventricle mask was dilated by 8 mm and applied to the WMH labels to calculate PV WMH and deep WMH volumes, i.e., all voxels inside the dilated ventricular mask were taken as PV WMH, and all remaining voxels outside the dilated ventricular mask were taken as deep WMH.

### Vascular Composite

Because certain elements necessary to compute the Framingham Score were not obtained in ADNI, we developed a vascular composite (VC) score to examine whether vascular risk factors, when combined together, influenced WMH load in males and females. The VC is a sum of questions regarding vascular conditions, diabetes, 0-1; alcohol abuse, 0-1; smoking history, 0-1; high systolic blood pressure, 0-1; high diastolic blood pressure, 0-1; and high BMI, 0-1; where 0 represents no and 1 represents yes, as well as Hachinski score, 0-4. The VC score was used as a continuous measure ranging from 0 (endorsing no conditions) to 10 (endorsing all conditions and maximum Hachinski score).

### Statistical Analysis

Independent sample t-tests and chi-square analysis were completed on demographic and clinical information using MATLAB R2019b. Bonferroni correction was used to account for multiple comparisons on the demographic and clinical information. WMH volumes were log-transformed to achieve a more normal distribution. Linear mixed effects models were used to investigate the association between WMH load (whole brain and subregions: frontal, temporal, parietal, occipital, deep, and periventricular) and vascular risk factors and each sex independently. Left and right lobes were summed for regional WMH values. All continuous values (including log-transformed WMH volumes) were z-scored within the population prior to the analyses. All p-values are reported as raw values with significance determined by false discovery rate FDR correction at 0.05^37^.

The first analysis was completed to determine if WMH progression rates differ between males and females. Baseline diagnosis, education, APOE4 status (defined as either 0 or 1; 0 = no □4 alleles and 1 = 1 or 2 □4 alleles), sex, and age were included as covariates. The interaction of interest was Sex:Age, contrasting rate of change with increased age in males against females. The model was run separately for WMH load (total, then frontal, temporal, parietal, occipital, and also deep and PV).

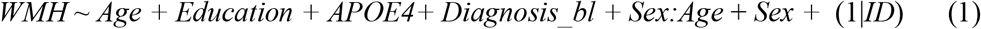

For the next set of analyses, the VC score was the variable of interest to determine whether vascular risk factors influenced WMH in males and females. The mixed effects model also included age, education, APOE4 status, and baseline diagnosis (contrasting MCI and AD against normal controls; NC), as covariates. Participant ID was included as a categorical random effect. The model was run separately for each WMH load (total, then frontal, temporal, parietal, occipital, and also deep and PV).

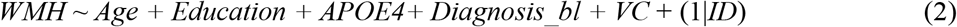

A follow-up model was used to determine whether the VC interacted with sex to influence WMH burden in males vs. females. The interaction of interest was VC:Sex (contrasting WMH burden in relation to the vascular composite for males against females). The model was run separately for each WMH load (total and then frontal, temporal, parietal, occipital and also deep and PV).

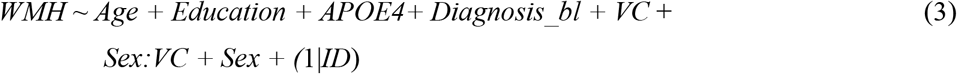

A second set of analyses was run to examine the influence of the individual vascular risk factors on WMH load. The model was run separately for males and females for each WMH load (total and then frontal, temporal, parietal, occipital, and also deep and PV).

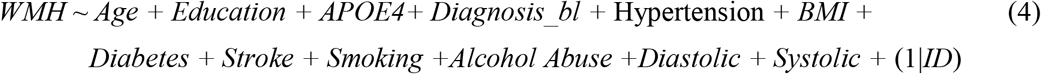

To determine whether the significant vascular risk factors interact with sex to influence WMH, a secondary level analysis was completed using linear mixed effects and including the significant risk factors from the fourth model (i.e., equation 4). The interactions of interest were Systolic:Sex and Hypertension:Sex (contrasting WMH burden in relation to the risk factors for males against females) in eq. (5). The model also included age, education, APOE4 status, and baseline diagnosis (contrasting MCI and AD against NC) as covariates. Participant ID was included as a categorical random effect. The model was run separately for each WMH load (total, then frontal, temporal, parietal, occipital, and also deep and PV).

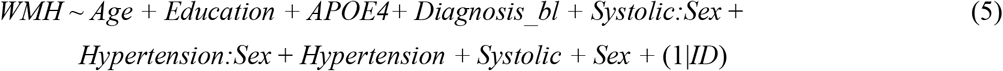

A final analysis was completed using linear mixed effects to examine whether there was an association between sex and WMH load on cognition (for three tests: global cognition, memory, and executive functioning). The interaction of interest was WMH:Sex (contrasting the difference in cognition in relation to WMH for males against females). The model also included age, education, and APOE4 status, baseline diagnosis (contrasting MCI and AD against NC), and VC as covariates. Participant ID was included as a categorical random effect. The models were run separately for each WMH load (total, then frontal, temporal, parietal, and occipital, and also deep and PV).

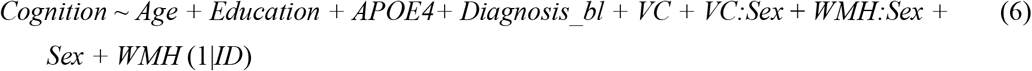

## Results

### Demographics

Table 1 presents demographic and clinical characteristics of study participants. Males were older than females (74.2y vs. 72.3y; *t*=6.24, *p*<.001) and had a higher education (16.4y vs. 15.6y; *t*=7.46, *p*<.001). Males had higher percentage of people with diabetes (10% vs 5% participants, x^2^=13.19, *p*<.001), history of hypertension (51% vs 44%, x^2^=9.03, *p*=.002), history of smoking (39% vs 28%, x^2^=25.85, *p*<.001), and alcohol abuse (6% vs 2%, x^2^=14.21, *p*<.001) than females. When looking at diagnostic status, men had a higher percentage of MCI participants (40% vs. 31%, x^2^=11.09, *p*<.001) and fewer cognitively healthy older adults (42% vs 54%, x^2^=34.91, *p*<.001). No other demographic or clinical features remained statistically significant after correction for multiple comparison.

**Table 1:**
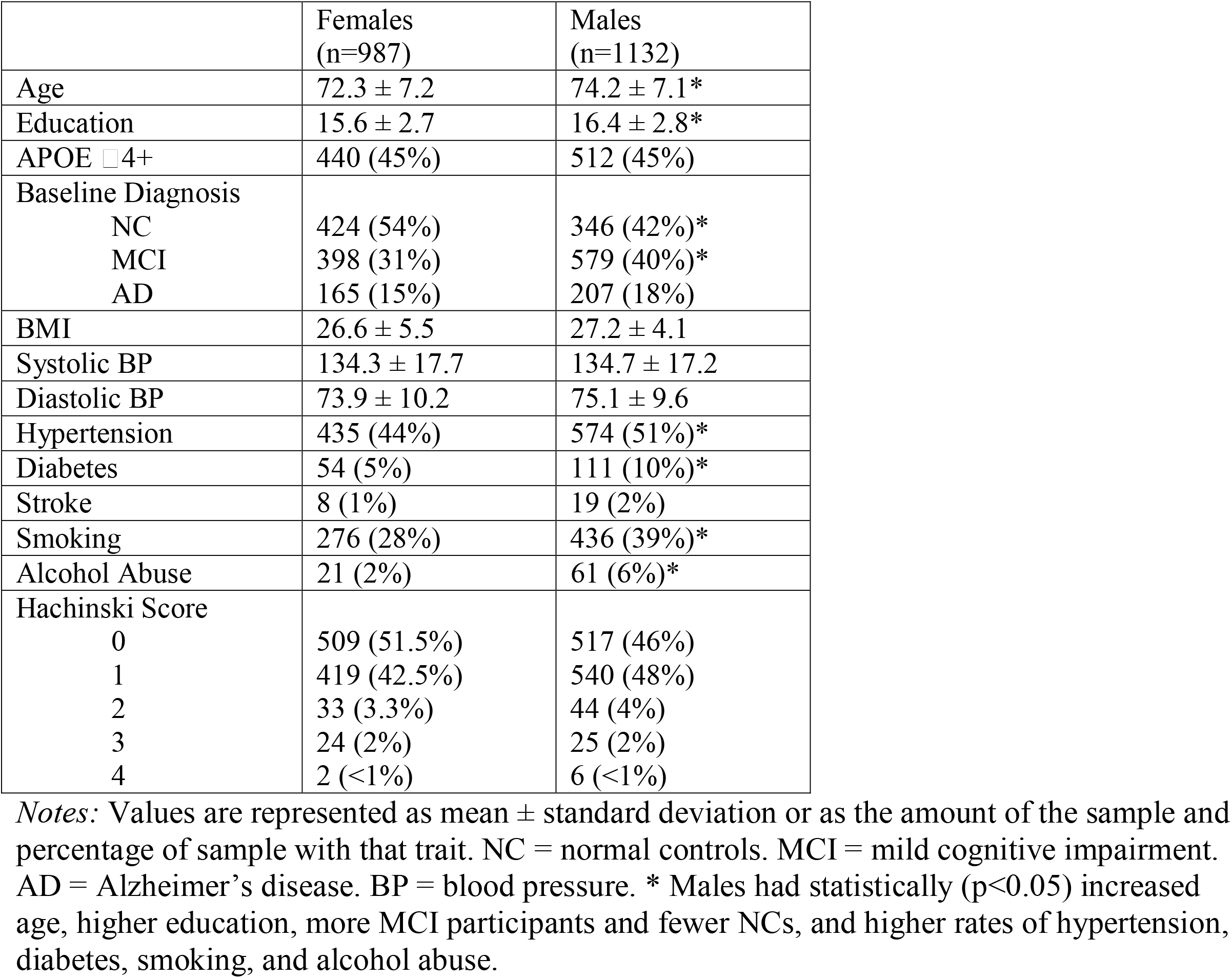
Demographic and clinical information for study participants.

|  | Females<br>(n=987) | Males<br>(n=1132) |
| --- | --- | --- |
| Age | 72.3 ± 7.2 | 74.2 ± 7.1* |
| Education | 15.6 ± 2.7 | 16.4 ± 2.8* |
| APOE ε4+ | 440 (45%) | 512 (45%) |
| Baseline Diagnosis |  |  |
| NC | 424 (54%) | 346 (42%)* |
| MCI | 398 (31%) | 579 (40%)* |
| AD | 165 (15%) | 207 (18%) |
| BMI | 26.6 ± 5.5 | 27.2 ± 4.1 |
| Systolic BP | 134.3 ± 17.7 | 134.7 ± 17.2 |
| Diastolic BP | 73.9 ± 10.2 | 75.1 ± 9.6 |
| Hypertension | 435 (44%) | 574 (51%)* |
| Diabetes | 54 (5%) | 111 (10%)* |
| Stroke | 8 (1%) | 19 (2%) |
| Smoking | 276 (28%) | 436 (39%)* |
| Alcohol Abuse | 21 (2%) | 61 (6%)* |
| Hachinski Score |  |  |
| 0 | 509 (51.5%) | 517 (46%) |
| 1 | 419 (42.5%) | 540 (48%) |
| 2 | 33 (3.3%) | 44 (4%) |
| 3 | 24 (2%) | 25 (2%) |
| 4 | 2 (<1%) | 6 (<1%) |

*Notes:* Values are represented as mean ± standard deviation or as the amount of the sample and percentage of sample with that trait. NC = normal controls. MCI = mild cognitive impairment. AD = Alzheimer's disease. BP = blood pressure. \* Males had statistically ( $p < 0.05$ ) increased age, higher education, more MCI participants and fewer NCs, and higher rates of hypertension, diabetes, smoking, and alcohol abuse.

### Progression of WMHs

WMH progression between males and females differed in all regions and measures except parietal (eq. 1). In the frontal (β = -0.07, SE = 0.02, *t =* -4.35, *p*<.001) region males had lower WMH progression with increased age compared to females. Males also had lower WMH progression in total (β = -0.03, SE = 0.01, *t =* -2.38, *p*=.01) and deep (β = -0.12, SE = 0.03, *t =* - 4.38, *p*<.001) brain. On the other hand, males had increased progression of WMHs in the occipital region (β = 0.11, SE = 0.02, *t =* 5.26, *p*<.001). Neither the temporal and parietal region analysis nor the periventricular region revealed sex differences in WMH progression with increased age.

### Vascular composite on WMHs

Table 2 presents the results from the linear mixed effects model examining the influence of the vascular composite score on male and females separately (eq. 2), as well as the interaction model (eq. 3). For males, age (*t* belongs to [14.68 – 61.21], *p*<.001) was significantly associated with increased WMHs in all regions except deep WMH. People with MCI had increased WMH load compared to NCs in all regions except deep WMH load (*t* belongs to [2.71 – 4.82], *p*<.006). People with AD had increased WMH compared to NCs in all regions and measures except deep WMH load (*t* belongs to [4.90 – 6.05], *p*<.001). Further, APOE positivity was associated with increased WMH burden in the parietal (*t* =2.49, *p*=.01) and occipital regions (*t* =2.90, *p*=.004). Importantly, an increased vascular composite score was associated with increased total, frontal, temporal, and parietal WMH burden (*t* belongs to [2.69 – 3.76], *p*<.007). That is, increased rates of vascular risk factors in males are associated with increased WMH accumulation in total, frontal, temporal, and parietal regions.

**Table 2:** Linear mixed effects examining vascular composite score influence on WMHs in males and females

| **Males (eq. 2)** | **Total** | **Frontal** | **Temporal** | **Parietal** | **Occipital** | **Deep** | **Periventricular** |
| --- | --- | --- | --- | --- | --- | --- | --- |
| Age | $\beta = 0.60$,<br>SE = 0.01,<br>$t = 61.21$,<br>$p < .001$ | $\beta = 0.56$,<br>SE < 0.01,<br>$t = 58.06$,<br>$p < .001$ | $\beta = 0.45$,<br>SE = .01,<br>$t = 39.71$,<br>$p < .001$ | $\beta = 0.60$,<br>SE = .01,<br>$t = 58.42$,<br>$p < .001$ | $\beta = 0.46$,<br>SE = .01,<br>$t = 35.02$,<br>$p < .001$ | $\beta = 0.03$,<br>SE = .02,<br>$t = 1.31$,<br>$p = 0.19$ | $\beta = 0.29$,<br>SE = .02,<br>$t = 14.68$,<br>$p < .001$ |
| MCI | $\beta = 0.24$,<br>SE = 0.06,<br>$t = 4.03$,<br>$p < .001$ | $\beta = 0.27$,<br>SE = 0.06,<br>$t = 4.43$,<br>$p < .001$ | $\beta = 0.17$,<br>SE = 0.06,<br>$t = 2.71$,<br>$p = .006$ | $\beta = 0.20$,<br>SE = .06,<br>$t = 3.31$,<br>$p < .001$ | $\beta = 0.18$,<br>SE = 0.07,<br>$t = 2.77$,<br>$p = .006$ | $\beta = -0.06$,<br>SE = 0.06,<br>$t = -0.97$,<br>$p = .33$ | $\beta = 0.25$,<br>SE = 0.05,<br>$t = 4.82$,<br>$p < .001$ |
| AD | $\beta = 0.48$,<br>SE = 0.08,<br>$t = 5.98$,<br>$p < .001$ | $\beta = 0.49$,<br>SE = 0.08,<br>$t = 6.05$,<br>$p < .001$ | $\beta = 0.38$,<br>SE = 0.09,<br>$t = 4.65$,<br>$p < .001$ | $\beta = 0.44$,<br>SE = 0.08,<br>$t = 5.52$,<br>$p < .001$ | $\beta = 0.43$,<br>SE = 0.09,<br>$t = 4.90$,<br>$p < .001$ | $\beta = -0.05$,<br>SE = 0.09,<br>$t = -0.53$,<br>$p = .60$ | $\beta = 0.37$,<br>SE = 0.07,<br>$t = 5.28$,<br>$p < .001$ |
| APOE $\epsilon 4+$ | $\beta = 0.11$,<br>SE = 0.05,<br>$t = 2.10$,<br>$p = .036$ | $\beta = 0.07$,<br>SE = 0.05,<br>$t = 1.30$,<br>$p = .19$ | $\beta = 0.10$,<br>SE = 0.05,<br>$t = 1.91$,<br>$p = .06$ | $\beta = 0.13$,<br>SE = 0.05,<br>$t = 2.49$,<br>$p = .01$ | $\beta = 0.17$,<br>SE = 0.06,<br>$t = 2.90$,<br>$p = .004$ | $\beta = 0.03$,<br>SE = 0.06,<br>$t = 0.48$,<br>$p = .63$ | $\beta = 0.06$,<br>SE = 0.05,<br>$t = 1.35$,<br>$p = .18$ |
| Education | $\beta = -0.03$,<br>SE = 0.03,<br>$t = -1.09$,<br>$p = .28$ | $\beta = -0.02$,<br>SE = 0.03,<br>$t = -0.79$,<br>$p = .43$ | $\beta = -0.02$,<br>SE = 0.03,<br>$t = -0.92$,<br>$p = .36$ | $\beta = -0.04$,<br>SE = 0.03,<br>$t = -1.48$,<br>$p = .14$ | $\beta = 0.02$,<br>SE = 0.03,<br>$t = 0.66$,<br>$p = .51$ | $\beta = -0.02$,<br>SE = 0.03,<br>$t = -0.84$,<br>$p = .40$ | $\beta < 0.01$,<br>SE = 0.03,<br>$t = 0.38$,<br>$p = .70$ |
| VC | $\beta = 0.02$,<br>SE < 0.01,<br>$t = 3.63$,<br>$p < .001$ | $\beta = 0.02$,<br>SE < 0.01,<br>$t = 3.76$,<br>$p < .001$ | $\beta = 0.02$,<br>SE < 0.01,<br>$t = 2.69$,<br>$p = .007$ | $\beta = 0.02$,<br>SE < 0.01,<br>$t = 3.20$,<br>$p = .001$ | $\beta = 0.01$,<br>SE < 0.01,<br>$t = 1.41$,<br>$p = .16$ | $\beta = 0.03$,<br>SE = 0.02,<br>$t = 2.06$,<br>$p = .039$ | $\beta = 0.34$,<br>SE = 0.02,<br>$t = 2.00$,<br>$p = .046$ |
| **Females (eq. 2)** | **Total** | **Frontal** | **Temporal** | **Parietal** | **Occipital** | **Deep** | **Periventricular** |
| Age | $\beta = 0.62$,<br>SE = 0.01,<br>$t = 51.04$,<br>$p < .001$ | $\beta = 0.61$,<br>SE = .01,<br>$t = 49.65$,<br>$p < .001$ | $\beta = 0.47$,<br>SE = .01,<br>$t = 33.81$,<br>$p < .001$ | $\beta = 0.60$,<br>SE = .01,<br>$t = 50.98$,<br>$p < .001$ | $\beta = 0.31$,<br>SE = .02,<br>$t = 16.96$,<br>$p < .001$ | $\beta = 0.18$,<br>SE = .02,<br>$t = 9.11$,<br>$p < .001$ | $\beta = 0.36$,<br>SE = .02,<br>$t = 16.41$,<br>$p < .001$ |
| MCI | $\beta = 0.21$, | $\beta = 0.19$, | $\beta = 0.21$, | $\beta = 0.22$, | $\beta = 0.13$, | $\beta = 0.04$, | $\beta = 0.20$, |
|  | **SE = 0.06,**<br>***t* = 3.47,**<br>***p* < .001** | **SE = 0.06,**<br>***t* = 3.17**<br>***p* = .001** | **SE = 0.06,**<br>***t* = 3.43**<br>***p* < .001** | **SE = 0.06,**<br>***t* = 3.60**<br>***p* < .001** | SE = 0.08,<br>*t* = 1.63<br>*p* = .10 | SE = 0.06,<br>*t* = 0.33<br>*p* = .51 | **SE = 0.05,**<br>***t* = 3.74**<br>***p* < .001** |
| AD | **$\beta$ = 0.40,**<br>**SE = 0.08,**<br>***t* = 4.91,**<br>***p* < .001** | **$\beta$ = 0.43,**<br>**SE = 0.08,**<br>***t* = 5.18,**<br>***p* < .001** | **$\beta$ = 0.35,**<br>**SE = 0.08,**<br>***t* = 4.17,**<br>***p* < .001** | **$\beta$ = 0.37,**<br>**SE = 0.08,**<br>***t* = 4.38,**<br>***p* < .001** | **$\beta$ = 0.17,**<br>**SE = 0.11,**<br>***t* = 2.88,**<br>***p* = .004** | $\beta$ = -0.20,<br>SE = 0.09,<br>*t* = -0.18<br>*p* = .86 | **$\beta$ = 0.44,**<br>**SE = 0.07,**<br>***t* = 5.94**<br>***p* < .001** |
| APOE ε4+ | $\beta$ = 0.04,<br>SE = 0.06,<br>*t* = 0.73,<br>*p* = .46 | $\beta$ = 0.02,<br>SE = 0.06,<br>*t* = 0.31,<br>*p* = .76 | $\beta$ = 0.05,<br>SE = 0.06,<br>*t* = 0.93,<br>*p* = .35 | $\beta$ = 0.04,<br>SE = 0.06,<br>*t* = 0.79,<br>*p* = .43 | **$\beta$ = 0.17,**<br>**SE = 0.06,**<br>***t* = 2.88,**<br>***p* = .004** | $\beta$ < -0.06,<br>SE = 0.06,<br>*t* = -1.08,<br>*p* = .28 | $\beta$ = 0.02,<br>SE = 0.05,<br>*t* = 0.46,<br>*p* = .64 |
| Education | $\beta$ = 0.03,<br>SE = 0.03,<br>*t* = 0.89,<br>*p* = .37 | $\beta$ = 0.02,<br>SE = 0.03,<br>*t* = 0.72,<br>*p* = .47 | $\beta$ = 0.03,<br>SE = 0.03,<br>*t* = 1.08,<br>*p* = .28 | $\beta$ = 0.03,<br>SE = 0.03,<br>*t* = 0.93<br>*p* = .35 | **$\beta$ = 0.08,**<br>**SE = 0.04,**<br>***t* = 2.28**<br>***p* = .022** | $\beta$ = -0.05,<br>SE = 0.03,<br>*t* = 1.57<br>*p* = .11 | $\beta$ = 0.06,<br>SE = 0.03,<br>*t* = 2.45,<br>*p* = .014 |
| VC | **$\beta$ = 0.04,**<br>**SE < 0.01,**<br>***t* = 4.05,**<br>***p* < .001** | **$\beta$ = 0.04,**<br>**SE < 0.01,**<br>***t* = 3.79,**<br>***p* < .001** | **$\beta$ = 0.04,**<br>**SE = 0.01,**<br>***t* = 3.49,**<br>***p* < .001** | **$\beta$ = 0.03,**<br>**SE < 0.01,**<br>***t* = 2.80,**<br>***p* = .005** | **$\beta$ = 0.04,**<br>**SE = 0.01,**<br>***t* = 3.15,**<br>***p* = .002** | **$\beta$ = 0.04,**<br>**SE = 0.03,**<br>***t* = 2.68,**<br>***p* = .007** | **$\beta$ = 0.05,**<br>**SE = 0.02,**<br>***t* = 2.73**<br>***p* = .006** |
| **Interaction**<br>**Model (eq. 3)** | **Total** | **Frontal** | **Temporal** | **Parietal** | **Occipital** | **Deep** | **Periventricular** |
| Male Sex | $\beta$ = -0.04,<br>SE = 0.04,<br>*t* = -1.05,<br>*p* = .29 | **$\beta$ = -0.14,**<br>**SE = 0.04,**<br>***t* = -3.57**<br>***p* < .001** | **$\beta$ = 0.11,**<br>**SE = 0.04,**<br>***t* = 2.79,**<br>***p* = .005** | $\beta$ = 0.03,<br>SE = .04,<br>*t* = 0.73,<br>*p* = .46 | **$\beta$ = 0.22,**<br>**SE = 0.04,**<br>***t* = 5.23,**<br>***p* < .001** | **$\beta$ = -0.19,**<br>**SE = 0.05,**<br>***t* = -3.58,**<br>***p* < .001** | $\beta$ < 0.01,<br>SE = 0.03,<br>*t* = 0.11,<br>*p* = .92 |
| MaleSex*VC | $\beta$ = -0.01,<br>SE = 0.01,<br>*t* = -0.83<br>*p* = .40 | $\beta$ = -0.01,<br>SE = 0.01,<br>*t* = -0.52,<br>*p* = .60 | $\beta$ = -0.01,<br>SE = 0.01,<br>*t* = -1.03,<br>*p* = .30 | $\beta$ < -0.01,<br>SE = 0.01,<br>*t* = -0.05,<br>*p* = .96 | $\beta$ = -0.02,<br>SE = 0.02,<br>*t* = -1.40,<br>*p* = .16 | $\beta$ = -0.01,<br>SE = 0.03,<br>*t* = -0.48,<br>*p* = .63 | $\beta$ = -0.02,<br>SE = 0.03,<br>*t* = -0.71,<br>*p* = .48 |

For females, age (*t* belongs to [9.11 – 51.04], *p*<.001) was significantly associated with increased WMHs in all regions. People with MCI had increased WMH load compared to NCs in all regions except occipital and deep WMH load (*t* belongs to [3.17 – 3.74], *p*<.001). People with AD had increased WMH compared to NCs in all regions and measures except deep WMH load (*t* belongs to [2.88 – 5.94], *p*<.005). APOE status was associated with increased WMH burden in only the occipital region (*t* =2.88, *p*<.004). Importantly, an increased vascular composite score was associated with increased WMH burden at all regions and measures (*t* belongs to [2.50 – 4.05], *p*<.05). This finding indicates that increased rates of vascular risk factors are associated with a widespread accumulation of WMHs in females in all regions and measures.

### Interaction of vascular composite with sex on WMHs

The results of interest for the model in eq. 3 included the main effect of Sex and the Sex:VC interaction. This interaction will provide information on whether the vascular component influences rate of WMH change over time differently in men and women. To avoid repetition with the previous section main effects of age and diagnosis will not be reported. In this model male sex was significantly associated with lower WMH accumulation over time in the frontal region (*t*= -3.57, *p*<.001) and deep regions (*t*= -3.58, *p*<.001) but with increased WMH accumulation over time in the temporal (*t*=2.79, *p*=.005) and occipital regions (*t*= 5.23, *p<*.001) in contrast to females. The interaction Sex:VC was not significant in any region. That is, the vascular composite score was not associated with increased rate of change in females vs. males.

### Individual risk factors on WMHs

Table 3 presents the output for the independent association for each individual risk factor on WMH burden in males and females separately (eq. 4). Only risk factors that showed significant associations with either group are presented in the table. Neither BMI, diastolic blood pressure, diabetes, smoking tobacco, nor alcohol abuse were significantly associated with WMH load in males or females. Systolic blood pressure and hypertension were associated with WMH load in both males and females. More specifically, increased systolic blood pressure was associated with total, frontal, and occipital WMHs in males (*t* belongs to [2.14 – 2.69], *p*<.05). History of hypertension was associated with increased total, frontal, temporal, parietal, deep, and periventricular WMHs in males (*t* belongs to [3.19 – 6.09], *p*<.005). In females, systolic blood pressure was associated with only deep (*t*=2.75, *p*=.005) and periventricular (*t*=2.25, *p*=0.24) WMHs, while history of hypertension was associated with increased frontal, deep, and periventricular WMHs (*t* belongs to [2.44 – 2.72], *p*<.05).

**Table 3:** Linear mixed effects examining the influence of independent risk factors on WMHs in males and females

|  |  |  |  |  |  |  |  |
| --- | --- | --- | --- | --- | --- | --- | --- |
| **Males (eq. 4)** | **Total** | **Frontal** | **Temporal** | **Parietal** | **Occipital** | **Deep** | **Periventricular** |
| Systolic BP | $\beta = 0.01$,<br>SE < 0.01,<br>$t = 2.32$,<br>$p = .020$ | $\beta < 0.01$,<br>SE < 0.01,<br>$t = 2.14$,<br>$p = .032$ | $\beta < 0.01$,<br>SE < 0.01,<br>$t = 1.09$,<br>$p = .27$ | $\beta = 0.01$,<br>SE < 0.01,<br>$t = 1.71$,<br>$p = .09$ | $\beta = 0.02$,<br>SE < 0.01,<br>$t = 2.69$,<br>$p = .007$ | $\beta < -0.01$,<br>SE = 0.01,<br>$t = -0.49$,<br>$p = .63$ | $\beta = 0.01$,<br>SE = 0.02,<br>$t = 0.81$,<br>$p = .42$ |
| History Of Hypertension | $\beta = 0.27$,<br>SE = 0.05,<br>$t = 5.18$,<br>$p < .001$ | $\beta = 0.32$,<br>SE = 0.05,<br>$t = 6.09$,<br>$p < .001$ | $\beta = 0.17$,<br>SE = 0.05,<br>$t = 3.20$,<br>$p = .001$ | $\beta = 0.23$,<br>SE = 0.06,<br>$t = 3.19$,<br>$p = .001$ | $\beta < 0.01$,<br>SE = 0.06,<br>$t = 0.13$,<br>$p = .89$ | $\beta = 0.19$,<br>SE = 0.06,<br>$t = 3.19$,<br>$p = .001$ | $\beta = 0.15$,<br>SE = 0.05,<br>$t = 3.26$,<br>$p = .001$ |
| **Females (eq. 4)** | **Total** | **Frontal** | **Temporal** | **Parietal** | **Occipital** | **Deep** | **Periventricular** |
| Systolic blood pressure | $\beta < 0.01$,<br>SE < 0.01,<br>$t = 0.95$,<br>$p = .34$ | $\beta < 0.01$,<br>SE < 0.01,<br>$t = 0.43$,<br>$p = .67$ | $\beta < 0.01$,<br>SE < 0.01,<br>$t = 1.19$,<br>$p = .23$ | $\beta < 0.01$,<br>SE < 0.01,<br>$t = 1.09$,<br>$p = .27$ | $\beta = 0.01$,<br>SE < 0.01,<br>$t = 0.20$,<br>$p = .84$ | $\beta = 0.03$,<br>SE = 0.01,<br>$t = 2.75$,<br>$p = .005$ | $\beta = 0.03$,<br>SE = 0.02,<br>$t = 2.25$,<br>$p = .024$ |
| History of Hypertension | $\beta = 0.12$,<br>SE = 0.06,<br>$t = 2.10$,<br>$p = .037$ | $\beta = 0.14$,<br>SE = 0.06,<br>$t = 2.44$,<br>$p = .015$ | $\beta = 0.04$,<br>SE = 0.06,<br>$t = 0.70$,<br>$p = .49$ | $\beta = 0.10$,<br>SE = 0.06,<br>$t = 1.82$,<br>$p = .07$ | $\beta = -0.02$,<br>SE = 0.01,<br>$t = -0.28$,<br>$p = .78$ | $\beta = 0.17$,<br>SE = 0.06,<br>$t = 2.92$,<br>$p = .003$ | $\beta = 0.14$,<br>SE = 0.05,<br>$t = 2.72$,<br>$p = .006$ |
| **Interaction Model (eq. 5)** | **Total** | **Frontal** | **Temporal** | **Parietal** | **Occipital** | **Deep** | **Periventricular** |
| MaleSex*SystolicBP | $\beta < 0.01$,<br>SE < 0.01,<br>$t = 1.06$,<br>$p = .29$ | $\beta < 0.01$,<br>SE < 0.01,<br>$t = 0.86$,<br>$p = .39$ | $\beta < 0.01$,<br>SE < 0.01,<br>$t = 0.31$,<br>$p = .76$ | $\beta < 0.01$,<br>SE < 0.01,<br>$t = 1.19$,<br>$p = .24$ | $\beta = 0.02$,<br>SE < 0.01,<br>$t = 2.18$,<br>$p = .029$ | $\beta = -0.03$,<br>SE = 0.01,<br>$t = -2.23$,<br>$p = .026$ | $\beta = -0.02$,<br>SE = 0.02,<br>$t = -1.66$,<br>$p = .24$ |
| Sex:Hypertension | $\beta = 0.15$,<br>SE = 0.07,<br>$t = 1.94$,<br>$p = .052$ | $\beta = 0.22$,<br>SE = 0.09,<br>$t = 2.47$,<br>$p = .014$ | $\beta = 0.13$,<br>SE = 0.08,<br>$t = 1.65$,<br>$p = .09$ | $\beta = 0.12$,<br>SE = 0.08,<br>$t = 1.61$,<br>$p = .11$ | $\beta = 0.07$,<br>SE = 0.08,<br>$t = 0.82$,<br>$p = .41$ | $\beta < -0.01$,<br>SE = 0.01,<br>$t = -0.06$,<br>$p = .95$ | $\beta = 0.03$,<br>SE = 0.07,<br>$t = 0.38$,<br>$p = .70$ |

### Interaction of risk factors with sex on WMHs

The results of interest from model in eq. 5 include the interaction between Systolic:Sex and Hypertension:Sex. Males with a history of hypertension had increased WMH burden in the frontal region compared to females (*t*=2.47, *p*=.014) and males with a high systolic blood pressure had increased WMH burden in the occipital region (*t*=2.18, *p*=.029). On the other hand, females with high systolic blood pressure had increased rates of deep WMH burden compared to males (*t*=2.23, *p*=.026). To ensure that differences in demographics and diagnostic group distributions did not impact our results, the models were repeated matching participant groups by age and diagnostic status. Similar results were obtained when matching the samples based on age and diagnostic status.

### Cognitive outcomes

Table 4 presents the estimates from the linear mixed effects model for change in cognition associated with WMHs for males and females (eq. 6). Increased WMH burden was associated with higher values (i.e., worse global cognition) on the CDR-SB at all regions and measures (*t* belongs to [3.23–17.72], *p*<.001) for females. This finding demonstrates that increased WMH burden is associated with worse global cognition in females. The male sex by WMH interaction revealed that males had a less steep slope than females at all regions and measures except deep WMHs (*t* belongs to [-2.43 – -4.56], *p*<.02), indicating that females have increased change in global cognition compared to males that is associated with WMHs.

**Table 4:** Linear mixed effects model showing the interactions between cognition and WMH for females and males

|  |  |  |  |  |  |  |  |
| --- | --- | --- | --- | --- | --- | --- | --- |
|  | Total | Frontal | Temporal | Parietal | Occipital | Deep | Periventricular |
| **CDR-SB (eq. 6)** |  |  |  |  |  |  |  |
| WMH | $\beta = 0.34,$<br>$SE = 0.02,$<br>$t = 17.40,$<br>$p < .001$ | $\beta = 0.34,$<br>$SE = 0.02,$<br>$t = 17.72,$<br>$p < .001$ | $\beta = 0.19,$<br>$SE = 0.02,$<br>$t = 10.45,$<br>$p < .001$ | $\beta = 0.27,$<br>$SE = 0.02,$<br>$t = 14.18,$<br>$p < .001$ | $\beta = 0.24,$<br>$SE = 0.02,$<br>$t = 14.34,$<br>$p < .001$ | $\beta = 0.06,$<br>$SE = 0.02,$<br>$t = 3.23$<br>$p = .001$ | $\beta = 0.06,$<br>$SE = 0.02,$<br>$t = 5.25$<br>$p < .001$ |
| Male:WMH | $\beta = -0.11,$<br>$SE = 0.02,$<br>$t = -4.42,$<br>$p < .001$ | $\beta = -0.11,$<br>$SE = 0.02,$<br>$t = -4.56$<br>$p < .001$ | $\beta = -0.10,$<br>$SE = 0.02,$<br>$t = -4.12$<br>$p < .002$ | $\beta = -0.09,$<br>$SE = 0.02,$<br>$t = -3.62$<br>$p < .001$ | $\beta = -0.06,$<br>$SE = 0.02,$<br>$t = -2.43$<br>$p = .015$ | $\beta < 0.01,$<br>$SE = 0.02,$<br>$t = 0.22$<br>$p = .82$ | $\beta = -0.06,$<br>$SE = 0.02,$<br>$t = -3.56$<br>$p < .001$ |
| Male:VC | $\beta < -0.01,$<br>$SE = 0.02,$<br>$t = -0.15,$<br>$p = .88$ | $\beta < -0.01,$<br>$SE = 0.02,$<br>$t = -0.34,$<br>$p = .73$ | $\beta < -0.01,$<br>$SE = 0.02,$<br>$t = -0.19,$<br>$p = .85$ | $\beta < -0.01,$<br>$SE = 0.02,$<br>$t = -0.20,$<br>$p = .84$ | $\beta < -0.01,$<br>$SE = 0.02,$<br>$t = -0.27,$<br>$p = .79$ | $\beta < -0.01,$<br>$SE = 0.02,$<br>$t = -0.41,$<br>$p = .68$ | $\beta < -0.01,$<br>$SE = 0.02,$<br>$t = -0.42,$<br>$p = .68$ |
| **TMT-B** (eq. 6) |  |  |  |  |  |  |  |
| WMH | $\beta = 0.23,$<br>$SE = 0.02,$<br>$t = 10.02,$<br>$p < .001$ | $\beta = 0.20,$<br>$SE = 0.03,$<br>$t = 8.75,$<br>$p < .001$ | $\beta = 0.16,$<br>$SE = 0.02,$<br>$t = 7.32,$<br>$p < .001$ | $\beta = 0.22,$<br>$SE = 0.03,$<br>$t = 9.62,$<br>$p < .001$ | $\beta = 0.21,$<br>$SE = 0.02,$<br>$t = 9.22,$<br>$p < .001$ | $\beta = 0.07,$<br>$SE = 0.02,$<br>$t = 2.82$<br>$p = .005$ | $\beta = 0.04,$<br>$SE = 0.02,$<br>$t = 2.52$<br>$p = .012$ |
| Male:WMH | $\beta = -0.05,$<br>$SE = 0.03,$<br>$t = -0.02,$<br>$p = .98$ | $\beta = 0.02,$<br>$SE = 0.03,$<br>$t = 0.78,$<br>$p = .44$ | $\beta = -0.04,$<br>$SE = 0.03,$<br>$t = -1.25,$<br>$p = .21$ | $\beta = -0.03,$<br>$SE = 0.03,$<br>$t = -0.90,$<br>$p = .37$ | $\beta < -0.01,$<br>$SE = 0.02,$<br>$t = -0.04,$<br>$p = .97$ | $\beta = -0.07,$<br>$SE = 0.03,$<br>$t = -2.67$<br>$p = .007$ | $\beta = -0.01,$<br>$SE = 0.02,$<br>$t = -0.29$<br>$p = .77$ |
| Male:VC | $\beta = 0.02,$<br>$SE = 0.03,$<br>$t = 0.83,$<br>$p = .41$ | $\beta = 0.02,$<br>$SE = 0.03,$<br>$t = 0.61,$<br>$p = .54$ | $\beta = 0.03,$<br>$SE = 0.03,$<br>$t = 0.91,$<br>$p = .36$ | $\beta = 0.02,$<br>$SE = 0.03,$<br>$t = 0.87,$<br>$p = .38$ | $\beta = 0.03,$<br>$SE = 0.03,$<br>$t = 0.91,$<br>$p = .36$ | $\beta = 0.02,$<br>$SE = 0.03,$<br>$t = 0.67,$<br>$p = .50$ | $\beta = 0.02,$<br>$SE = 0.03,$<br>$t = 0.66,$<br>$p = .50$ |
| **RAVLT- Immediate**<br>(eq. 6) |  |  |  |  |  |  |  |
| WMH | $\beta = -0.24,$<br>$SE = 0.02,$<br>$t = -12.15,$<br>$p < .001$ | $\beta = -0.22,$<br>$SE = 0.02,$<br>$t = -11.64,$<br>$p < .001$ | $\beta = -0.15,$<br>$SE = 0.02,$<br>$t = -8.17,$<br>$p < .001$ | $\beta = -0.22,$<br>$SE = 0.02,$<br>$t = -11.07$<br>$p < .001$ | $\beta = -0.14,$<br>$SE = 0.02,$<br>$t = -8.89,$<br>$p < .001$ | $\beta = 0.02,$<br>$SE = 0.02,$<br>$t = 1.21$<br>$p = .22$ | $\beta = -0.04,$<br>$SE = 0.01,$<br>$t = -3.59$<br>$p < .001$ |
| Male:WMH | $\beta = 0.06,$ | $\beta = 0.06,$ | $\beta = 0.04,$ | $\beta = 0.06,$ | $\beta = 0.02,$ | $\beta = -0.02,$ | $\beta = 0.03,$ |
|  | **SE = 0.02,**<br>***t* = 2.55**<br>***p* = .01** | **SE = 0.02,**<br>***t* = 2.63,**<br>***p* = .008** | SE = 0.02,<br>*t* = 1.79,<br>*p* = .07 | **SE = 0.02,**<br>***t* = 2.33,**<br>***p* = .02** | SE = 0.02<br>*t* = 1.00,<br>*p* = .31 | SE = 0.02,<br>*t* = -0.74<br>*p* = .46 | SE = 0.01,<br>*t* = 1.82<br>*p* = .07 |
| Male:VC | $\beta < 0.01$,<br>SE = 0.02,<br>*t* = 0.11,<br>*p* = .90 | $\beta < 0.01$,<br>SE = 0.02,<br>*t* = 0.22,<br>*p* = .82 | $\beta < 0.01$,<br>SE = 0.02,<br>*t* = 0.10,<br>*p* = .92 | $\beta < 0.01$,<br>SE = 0.02,<br>*t* = 0.13,<br>*p* = .89 | $\beta < 0.01$,<br>SE = 0.02,<br>*t* = 0.09,<br>*p* = .92 | $\beta < 0.01$,<br>SE = 0.02,<br>*t* = 0.36,<br>*p* = .72 | $\beta < 0.01$,<br>SE = 0.02,<br>*t* = 0.20,<br>*p* = .85 |
| **RAVLT- Learning**<br>(eq. 6) |  |  |  |  |  |  |  |
| WMH | **$\beta$ = -0.13,**<br>**SE = 0.02,**<br>***t* = -5.68,**<br>***p* < .001** | **$\beta$ = -0.11,**<br>**SE = 0.02,**<br>***t* = -4.99,**<br>***p* < .001** | **$\beta$ = -0.08,**<br>**SE = 0.02,**<br>***t* = -3.69,**<br>***p* < .001** | **$\beta$ = -0.13,**<br>**SE = 0.02,**<br>***t* = -6.00,**<br>***p* < .001** | **$\beta$ = -0.09,**<br>**SE = 0.02,**<br>***t* = -4.65,**<br>***p* < .001** | $\beta$ = 0.02,<br>SE = 0.02,<br>*t* = 0.95<br>*p* = .34 | **$\beta$ = -0.07,**<br>**SE = 0.02,**<br>***t* = -4.30**<br>***p* < .001** |
| Male:WMH | $\beta$ = 0.04,<br>SE = 0.03,<br>*t* = 1.57,<br>*p* = .11 | $\beta$ = 0.03,<br>SE = 0.03,<br>*t* = 1.23,<br>*p* = .22 | $\beta$ = 0.02,<br>SE = 0.03,<br>*t* = 0.73,<br>*p* = .47 | $\beta$ = 0.05,<br>SE = 0.03,<br>*t* = 1.88,<br>*p* = .06 | $\beta$ = 0.03,<br>SE = 0.03<br>*t* = 1.26,<br>*p* = .21 | $\beta$ = -0.03,<br>SE = 0.03,<br>*t* = -1.02<br>*p* = .30 | **$\beta$ = 0.07,**<br>**SE = 0.02,**<br>***t* = 3.47**<br>***p* < .001** |
| Male:VC | $\beta$ = -.05,<br>SE = 0.03,<br>*t* = -1.83,<br>*p* = .07 | $\beta$ = -.05,<br>SE = 0.03,<br>*t* = -1.72,<br>*p* = .08 | $\beta$ = -.05,<br>SE = 0.03,<br>*t* = -1.78,<br>*p* = .075 | $\beta$ = -.05,<br>SE = 0.03,<br>*t* = -1.89,<br>*p* = .058 | $\beta$ = -.05,<br>SE = 0.03,<br>*t* = -1.79,<br>*p* = .073 | $\beta$ = -.04,<br>SE = 0.03,<br>*t* = -1.54,<br>*p* = .12 | $\beta$ = -.05,<br>SE = 0.03,<br>*t* = -1.79,<br>*p* = .072 |
| **RAVLT- Perc_Forgetting**<br>(eq. 6) |  |  |  |  |  |  |  |
| WMH | **$\beta$ = 0.09,**<br>**SE = 0.02,**<br>***t* = 3.59,**<br>***p* < .001** | **$\beta$ = 0.09,**<br>**SE = 0.02,**<br>***t* = 3.55,**<br>***p* < .001** | $\beta$ = 0.04,<br>SE = 0.02,<br>*t* = 1.75,<br>*p* = .08 | **$\beta$ = 0.07,**<br>**SE = 0.02,**<br>***t* = 2.17,**<br>***p* = .003** | **$\beta$ = 0.06,**<br>**SE = 0.02,**<br>***t* = 2.73,**<br>***p* = .006** | $\beta < 0.01$,<br>SE = 0.02,<br>*t* = -0.01<br>*p* = .99 | $\beta$ = 0.01,<br>SE = 0.02,<br>*t* = 0.67<br>*p* = .51 |
| Male:WMH | $\beta$ = -0.03,<br>SE = 0.03,<br>*t* = -0.90,<br>*p* = .36 | $\beta$ = -0.03,<br>SE = 0.03,<br>*t* = -1.00,<br>*p* = .32 | $\beta$ = -0.01,<br>SE = 0.03,<br>*t* = -0.05,<br>*p* = .96 | $\beta$ = -0.01,<br>SE = 0.03,<br>*t* = -0.46,<br>*p* = .64 | $\beta$ = -0.02,<br>SE = 0.03,<br>*t* = -0.72,<br>*p* = .47 | $\beta$ = 0.03,<br>SE = 0.03,<br>*t* = 1.03<br>*p* = .30 | $\beta$ = -0.04,<br>SE = 0.02,<br>*t* = -1.58<br>*p* = .11 |
| Male:VC | $\beta < 0.01$,<br>SE = 0.03,<br>*t* = 0.17, | $\beta < 0.01$,<br>SE = 0.03,<br>*t* = 0.11, | $\beta < 0.01$,<br>SE = 0.03,<br>*t* = 0.17, | $\beta < 0.01$,<br>SE = 0.03,<br>*t* = 0.18, | $\beta < 0.01$,<br>SE = 0.03,<br>*t* = 0.20, | $\beta < -0.01$,<br>SE = 0.03,<br>*t* = -0.05, | $\beta < -0.01$,<br>SE = 0.03,<br>*t* = 0.16, |
| | $p = .86$ | $p = .92$ | $p = .87$ | $p = .86$ | $p = .84$ | $p = .96$ | $p = .87$ |
| **FAQ** (eq. 6) |  |  |  |  |  |  |  |
| WMH | $\beta = 0.33,$<br>$SE = 0.02,$<br>$t = 16.46,$<br>$p < .001$ | $\beta = 0.23,$<br>$SE = 0.02,$<br>$t = 16.13,$<br>$p < .001$ | $\beta = 0.20,$<br>$SE = 0.02,$<br>$t = 10.31,$<br>$p < .001$ | $\beta = 0.29,$<br>$SE = 0.02,$<br>$t = 14.10,$<br>$p < .001$ | $\beta = 0.24,$<br>$SE = 0.02,$<br>$t = 13.78,$<br>$p < .001$ | $\beta = -0.03,$<br>$SE = 0.02,$<br>$t = -1.69$<br>$p = .091$ | $\beta = 0.06,$<br>$SE = 0.01,$<br>$t = 4.82$<br>$p < .001$ |
| Male:WMH | $\beta = -0.06,$<br>$SE = 0.03,$<br>$t = -2.40,$<br>$p = .02$ | $\beta = -0.06,$<br>$SE = 0.03,$<br>$t = -1.98$<br>$p = .047$ | $\beta = -0.07,$<br>$SE = 0.02,$<br>$t = -2.96$<br>$p = .003$ | $\beta = -0.06,$<br>$SE = 0.03,$<br>$t = -2.46$<br>$p = .014$ | $\beta = -0.02,$<br>$SE = 0.02,$<br>$t = -0.72$<br>$p = .47$ | $\beta = -0.07,$<br>$SE = 0.02,$<br>$t = -3.21$<br>$p = .001$ | $\beta = -0.07,$<br>$SE = 0.02,$<br>$t = -4.43$<br>$p = .001$ |
| Male:VC | $\beta < 0.01,$<br>$SE = 0.02,$<br>$t = 0.24,$<br>$p = .81$ | $\beta = -.01,$<br>$SE = 0.02,$<br>$t = -0.46,$<br>$p = .64$ | $\beta < 0.01,$<br>$SE = 0.02,$<br>$t = 0.15,$<br>$p = .88$ | $\beta < 0.01,$<br>$SE = 0.02,$<br>$t = 0.24,$<br>$p = .81$ | $\beta < 0.01,$<br>$SE = 0.02,$<br>$t = 0.14,$<br>$p = .88$ | $\beta = -0.01,$<br>$SE = 0.02,$<br>$t = -0.46,$<br>$p = .65$ | $\beta < -0.01,$<br>$SE = 0.02,$<br>$t = -0.29,$<br>$p = .77$ |

Notes: WMH = white matter hyperintensity. CDR-SB = Clinical dementia rating- sum of boxes. RAVLT = Ray's Auditory Verbal Learning test. TMT-B = Trail Making Test-B. Perc-Forgetting = Percent forgetting. FAQ = Functional Activities Questionnaire

When examining executive functioning, as measured by the Trail-Making Test Part B (TMT-B), there was an association between WMHs and executive functioning scores in females. Increased TMT-B scores was associated with increased WMH burden in females at all regions and measures (*t* belongs to [2.52 – 10.02], *p*<.05). The male sex by WMH interaction revealed that males had a less steep slope than females at the occipital region (*t =* -2.67, *p*=.007), indicating that females have increased change in executive functioning compared to males that is associated with WMHs in those two regions.

The Rey Auditory Verbal Learning Test (RAVLT) was used to examine the association between memory and WMHs. Both RAVLT immediate (*t* belongs to [-3.59 – -12.15], *p*<.001) and learning (*t* belongs to [-3.69 – 5.68], *p*<.05) were significantly associated with WMHs in females at all regions and measures except deep WMHs. For RAVLT percent forgetting, a significant association between WMHs was observed in females for total, frontal, parietal and occipital WMHs (*t* belongs to [2.17 – 3.59], *p*<.01). That is, increased WMHs were associated with worse memory performance in females. The male sex by WMH interaction for RAVLT immediate score was significant for total, frontal, and temporal WMH burden (*t* belongs to [2.33 –2.63], *p*<.05). The interaction between male sex and WMHs was also significant for RAVLT learning and periventricular WMHs (*t*=3.47, *p*=.001). That is, males had a less steep slope compared to females, indicating that more WMH accumulation is needed (in males) for the same change in RAVLT immediate and learning score as females.

The functional activities questionnaire (FAQ), was examined to assess instrumental activities of daily living. The relationship between WMHs and the FAQ was significant for females at all regions except deep WMHs (*t* belongs to [4.82 – 16.46], *p*<.001). The male sex by WMH interaction revealed that males had a less steep slope than females in total, temporal, parietal, deep, and periventricular WMH progression (*t* belongs to [-2.40 – -4.43], *p*<.05). These findings suggest that for females higher scores (i.e., worse functional abilities) is impacted by increased WMHs, this relationship does not occur in males in total. Temporal, parietal, deep, and periventricular WMH progression.

## Discussion

Despite the large amount of research examining the influence of WMHs on cognitive decline, there are limited investigations directly assessing the influence of sex on these relationships. The current study was designed to examine if sex affects the association between WMH burden and vascular risk factors as well as the association between WMHs and cognition. The current study shows that history of hypertension and systolic blood pressure are the main independent factors associated with WMH burden in males. On the other hand, in females few associations are observed with independent factors, with the strongest (and most) associations between WMHs and the vascular composite score. Furthermore, we observed a strong association between WMH burden and cognition in females in all domains (global cognition, executive functioning, memory, and functional activities). In males, however, the slope of the association was less steep compared to females (for global cognition, executive functioning, some memory scores, and functional activities), indicating that more WMH accumulation is needed in males to see the same change in those three domains.

We observed that compared to females, males demonstrated similar, increased, and decreased WMH progression depending on the regions observed. These findings are consistent with previous reports indicating both increased^16–18^ and decreased^20^ WMH burden in females relative to males. While females showed similar parietal WMHs, they exhibited lower WMH progression in only the occipital region relative to males. Consistent with previous studies, females showed the largest WMH progression compared to males in deep^15,16^ and frontal^15^ WMH burden followed by total WMH burden. When examining the interaction between risk factors and sex, we observed that the vascular composite did not interact with sex to influence rate of WMH progression. However, when examining the risk factors independently both systolic blood pressure and hypertension influenced rate of WMH progression. Relative to females, males with history of hypertension exhibited increased WMH progression in the frontal region and males with high systolic blood pressure had increased WMH progression in the occipital region. Females with high systolic blood pressure showed increased deep WMH progression compared to males. Taken together, including regional differences and multiple vascular risk factors when examining WMH burden is essential to observe how sex influences WMHs.

Consistent with previous research, WMH accumulation was associated with lower cognitive performance^2,38,39^. More specifically, change in global cognition and executive functioning was associated with WMH burden at all regions and measures for females. Males had less change in global cognition that was associated with WMHs than females. With respect to executive functioning, males had a less steep slope in the parietal and occipital regions, indicating less cognitive change in those regions associated with WMH burden. Previous research has also observed that WMHs are associated with change in memory performance in NCs, MCI, and AD^40^. Similarly, we observed that lower memory performance (i.e., high scores in RAVLT percent forgetting, and lower scores in RAVLT immediate and learning) was associated with increased WMH accumulation in females. Males showed less change in RAVLT immediate scores associated with total, frontal, and parietal WMH burden than females. With respect to RAVLT learning, the only sex difference was periventricular WMH burden, with males showing less change associated with WMH load relative to females. No difference in RAVLT percent forgetting scores were observed between males and females. Taken together, immediate memory appears to be more strongly associated with WMH in females compared to males. This finding indicates that WMH progression is associated with short-term memory (i.e., RAVLT immediate) differently in males and females but the relationship between WMHs and long-term memory (i.e., RAVLT percent forgetting) is similar in males and females. Previous research has also found that WMH burden is also associated with lower functional capacity and physical health, particularly in women^15^. The current study supports this finding, with females showing a strong relationship between higher scores on the FAQ (i.e., lower ability to independently perform activities of daily living) and WMH burden. On the other hand, males had a less steep slope compared to females in total, frontal, parietal, and occipital regions. That is, functional performance was less affected by WMH accumulation in males than females.

There are a few limitations and strengths of the current study. Our sample had an average education of 16 years and was comprised mainly of white individuals (93% of the sample was white), which may reduce generalizability to more representative samples. A major strength of this study is the use of the imaging processing methods that have been extensively tested and validated for use in multi-center and multi-scanner studies. The WMH segmentation technique on T1w images has shown strong correlations to WMH measures obtained from T2w/PD and FLAIR images^41^, and have been previously used to successfully observe WMHs in healthy aging and AD^4,38^.

Many of the underlying causes of cerebrovascular disease are preventable and/or treatable. Identifying which factors are associated with WMH accumulation could, therefore, enable timely interventions to mitigate cognitive decline progression and conversion to dementia. The findings presented here show that a history hypertension is the main factor influencing WMH development in males whereas for females a combination of several factors (i.e., alcohol consumption, history of hypertension, smoking, high BMI, diabetes, alcohol consumption, and Hachinski score) have the largest impact on WMH accumulation. This accumulation of factors may explain why the females experienced more cognitive decline associated with the WMHs. Females who endorse these vascular risk factors and develop WMHs may require less deposition of other pathologies that occur with age and dementia to develop cognitive symptoms. From a clinical standpoint, these findings suggest that interventions should target hypertension in males but all vascular factors in women to reduce deterioration in cognitive functioning and risk of future cognitive decline. These findings can help improve the develop of interventions to slow cognitive decline and dementia by encouraging researchers and clinicians to examine the techniques males and females separately.

## Data Availability

All data produced are available online from the Alzheimer's Disease Neuroimaging Initiative (ADNI) database (adni.loni.usc.edu).

https://adni.loni.usc.edu/

## Acknowledgments

Data collection and sharing for this project was funded by the Alzheimer’s Disease Neuroimaging Initiative (ADNI) (National Institutes of Health Grant U01 AG024904) and DOD ADNI (Department of Defense award number W81XWH-12-2-0012). ADNI is funded by the National Institute on Aging, the National Institute of Biomedical Imaging and Bioengineering, and through generous contributions from the following: AbbVie, Alzheimer’s Association; Alzheimer’s Drug Discovery Foundation; Araclon Biotech; BioClinica, Inc.; Biogen; Bristol-Myers Squibb Company; CereSpir, Inc.; Cogstate; Eisai Inc.; Elan Pharmaceuticals, Inc.; Eli Lilly and Company; EuroImmun; F. Hoffmann-La Roche Ltd and its affiliated company Genentech, Inc.; Fujirebio; GE Healthcare; IXICO Ltd.; Janssen Alzheimer Immunotherapy Research & Development, LLC.; Johnson & Johnson Pharmaceutical Research & Development LLC.; Lumosity; Lundbeck; Merck & Co., Inc.; Meso Scale Diagnostics, LLC.; NeuroRx Research; Neurotrack Technologies; Novartis Pharmaceuticals Corporation; Pfizer Inc.; Piramal Imaging; Servier; Takeda Pharmaceutical Company; and Transition Therapeutics. The Canadian Institutes of Health Research is providing funds to support ADNI clinical sites in Canada. Private sector contributions are facilitated by the Foundation for the National Institutes of Health (www.fnih.org). The grantee organization is the Northern California Institute for Research and Education, and the study is coordinated by the Alzheimer’s Therapeutic Research Institute at the University of Southern California. ADNI data are disseminated by the Laboratory for Neuro Imaging at the University of Southern California.

## Notes

**Funding information** Alzheimer’s Disease Neuroimaging Initiative; This research was supported by a grant from the Canadian Institutes of Health Research.

**Financial Disclosures** Dr. Morrison is supported by a postdoctoral fellowship from Canadian Institutes of Health Research, Funding Reference Number: MFE-176608. Dr. Dadar reports receiving research funding from the Healthy Brains for Healthy Lives (HBHL), Alzheimer Society Research Program (ASRP), and Douglas Research Centre (DRC). Dr. Collins reports receiving research funding from Canadian Institutes of Health research, the Canadian National Science and Engineering Research Council, Brain Canada, the Weston Foundation, and the Famille Louise & André Charron.

**Conflict of Interest** The authors declare no competing interests

### Competing Interest Statement

The authors have declared no competing interest.

### Funding Statement

Dr. Morrison is supported by a postdoctoral fellowship from Canadian Institutes of Health Research, Funding Reference Number: MFE-176608.
Dr. Dadar reports receiving research funding from the Healthy Brains for Healthy Lives (HBHL), Alzheimer Society Research Program (ASRP), and Douglas Research Centre (DRC).
Dr. Collins reports receiving research funding from Canadian Institutes of Health research, the Canadian National Science and Engineering Research Council, Brain Canada, the Weston Foundation, and the Famille Louise & Andre Charron.

### Author Declarations

The study used only openly available human data that were originally located from the Alzheimer's Disease Neuroimaging Initiative (ADNI) database (adni.loni.usc.edu).

